# Socio-demographic determinants associated with *Blastocystis* infection in Arequipa, Peru

**DOI:** 10.1101/2020.06.07.20124941

**Authors:** Renzo S. Salazar-Sánchez, Kasandra Ascuña-Durand, Ricardo Cartillo-Neyra, Victor Vásquez-Huerta, Elí Martínez-Barrios, Jorge Ballón-Echegaray

**Affiliations:** Laboratorio de Microbiología Molecular, Facultad de Medicina, Universidad Nacional de San Agustín, Arequipa, Peru; Zoonotic Disease Research Lab, One Health Unit, School of Public Health and Administration, Universidad Peruana Cayetano Heredia, Lima, Peru; Department of Biostatistics, Epidemiology & Informatics, Perelman School of Medicine of the University of Pennsylvania, Philadelphia, PA 19104, USA; Departamento de Microbiología y Patología, Facultad de Medicina, Universidad Nacional de San Agustín, Arequipa, Peru

**Keywords:** *Blastocystis*, One Health, parasites, risk factors, social determinants

## Abstract

*Blastocystis* is one of the most common protozoa in the human gut and is a zoonotic parasite related to unhealthy living conditions. This parasite shows a broad distribution, unclear symptomatology, and undefined pathogenicity. In Peru, studies report the presence of *Blastocystis* in many regions, but the highest prevalence levels are reported in Arequipa. The aim of this study was to link *Blastocystis* infection with social determinants of health. We recruited and survey 232 participants from infected and uninfected homes. All samples were analyzed by direct microscopy and confirmed with methylene-stained stool smear. We found a human *Blastocystis* prevalence of 51.3% in the study sample. We also found statistical associations between *Blastocystis* infection and the use of alternative non-domiciliary water supplies as well as the use of latrine for body-waste disposal, suggesting these are risk factors for human *Blastocystis* infection.

## Introduction

Intestinal parasitic infections are one of the most common public health problems, affecting more than two million people around the world (1,2). They are mainly found in areas with poor health and sanitary conditions, limited access to safe drinking water, inadequate disposal of human feces (3,4), and low levels of access to health care facilities (5). Among intestinal parasitic infections, *Blastocystis* is the most common causative agent (6).

*Blastocystis* is a parasite with a worldwide distribution and is the most commonly isolated microorganism in parasitological surveys (6). Despite being so ubiquitous and being discovered more than a 100 years ago (7), little is known about their pathogenicity, genetic diversity, transmission dynamics (including zoonotic and zooanthroponotic transmission), therapeutic options and treatment efficacy (8,9). The role of *Blastocystis* in human disease remains controversial (10). Its presence in symptomatic and asymptomatic patients is difficult to explain; therefore, some report is as a pathogen while others regard it as a commensal (11,12).

Many risk factors are associated with *Blastocystis* acquisition including being a 6 or 7 year-old child (13), being male (14,15) and flooding of the home. The use of a latrine is also associated with *Blastocystis* acquisition likely because of its relation to poor hygiene (14,16), water-borne transmission, and the lack of access to treated drinking water (17,18). By contrast, other studies mentioned that sociodemographic factors such as age, gender, water quality, disposition of excreta, place of residence, number of children in the house, monthly income, type of property, floor type, wall type, availability of public services, hand washing habits and garbage disposal were not associated with *Blastocystis* infection (19). Domestic and wild animals are considered an important source of transmission of *Blastocystis* (8,20). However, previous studies suggest that livestock animals are not the main contributor of human infections (21). Carnivores, reptiles and insects also do not seem to be important sources of infection (20).

Most of these risk-factor studies have been conducted in Asia and Europe, with few studies focusing on Latin American countries. In Peru, the presence of *Blastocystis* has been reported in many regions, mainly in parasitological surveys carried out in school children (22–24). The most common association with *Blastocystis* infection reported among positive study sites was poor sanitary conditions (25,26). Among the 24 regions in Peru, Arequipa has reported the highest prevalence (25). These previous studies did not aim to find factors associated to the high prevalence of infection in Arequipa or evidence of their association with symptomatology and gastrointestinal illnesses in infected cases.

Due to *Blastocystis’* disputed role as a pathogenic agent and the scarcity of studies on factors associated with infection and symptomatology, this study aimed to identify individual and household level factors associated with *Blastocystis* infections, and to add new evidence for the understanding of the complex epidemiology of this controversial intestinal protozoon. This is the first attempt to study *Blastocystis* using the One Health approach in the region.

## Methods

### Ethical statement

The Institutional Review Boards of the Universidad Peruana Cayetano Heredia approved the protocol of this study, identification number 18006. Before collecting any data, all participants provided written informed consent. Minors provided verbal and/or written informed assent and their parents provided written informed consent. We included in the study participants from any age that were not taking any antimicrobial or antiparasitic treatment at the time of sampling collection or the prior seven days. All participants completed an epidemiological survey to assess clinical and sanitary living conditions. Participants’ questions were solved at any moment.

### Study site

This study was conducted in Arequipa city, which is located in the south highlands with a population size around a million people (27). Arequipa city comprises periurban and urban areas, which have distinct migration histories, and continuous growth from the center to the periphery (28). In the context of this centrifugal expansion, the periurban communities are younger than the urban communities and are farther from the center of the city (28). There are also social differences between the two types of communities: periurban areas tend to have limited basic services like electricity, water, sanitation, health and education, whereas urban locations have all these services more readily available (Figure 1).

**Figure 1.**
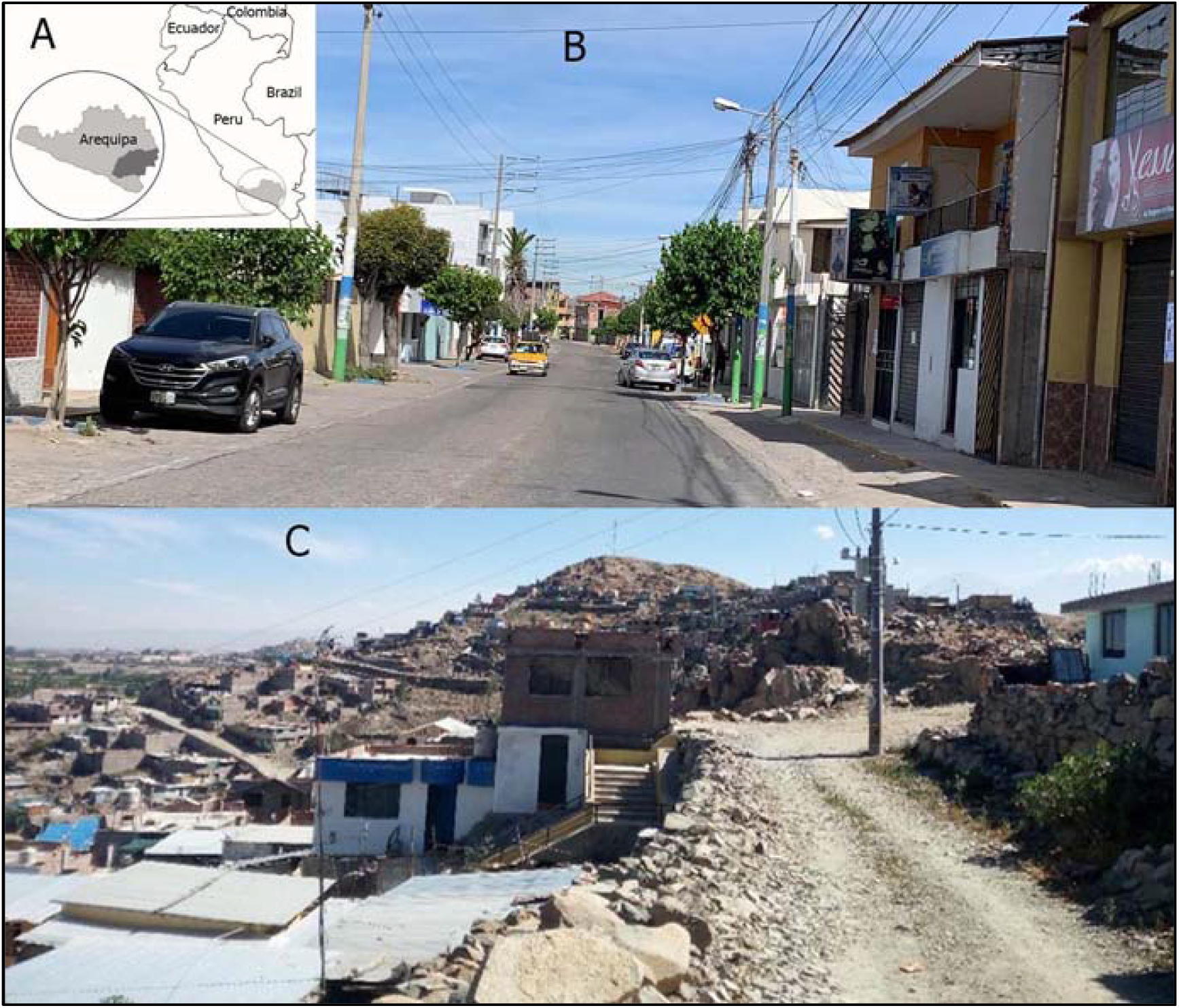
Study site: A) Map of Arequipa in Peru. B) Urban locality with adequate sanitary conditions and access to public facilities. C) Peri-urban locality with limited sanitary conditions.

### Study Population and participants selection

We carried out two free parasitological screening campaigns. The first campaign was advertised on the radio (Radio Universidad Arequipa) and television (TV UNSA) for three weeks in January, 2019, inviting any resident of Arequipa to participate in the campaign. The collection of samples, analysis and reporting of results occurred in the Molecular Microbiology Laboratory, Universidad Nacional de San Agustín de Arequipa in Peru between January and March of 2019. 168 people participated in the campaign and 61 of them agreed to participate in the study as index cases. The second campaign was focused on three elementary schools in periurban areas of Tiabaya district, Sachaca district (includes elementary and kinder garden level) and Socabaya district. Elementary school children in Peru are 6 to 11 years old. We coordinated a personal interview with the principal of each institution to ask for permission to carry out the campaign. Once they agreed to participate, we invited every student’s parent to ask their children to participate via letters or educational talks. Detailed information of participants of the campaigns and recruitment are included in figure 2.

**Figure 2.**
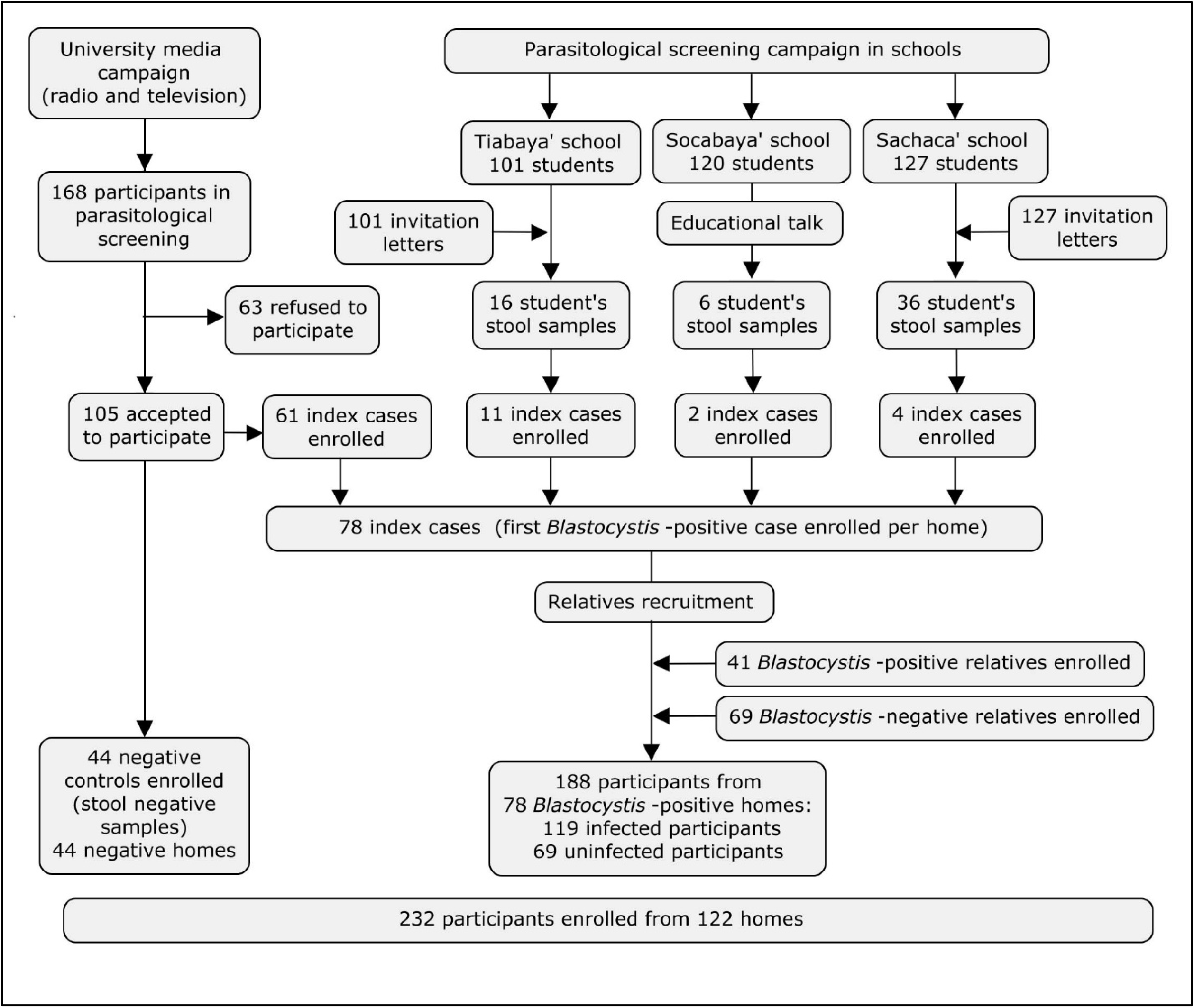
Diagram of participant’s distribution per home.

To recruit relatives of index cases, we communicated by phone or in person with index cases or parents from index cases to coordinate a date for visit their home. Then, we invited relatives and other household members to participate in the study with a free parasitological screening for them and their animals. All results were reported to participants in person two days after taking the stool sample. The research team medical doctor provided people who were infected by a different parasite besides *Blastocystis* with antiparasitic treatment.

### Sample collection and laboratory diagnosis

All samples were collected by each participant in a sterilized plastic wide mouth flask without additives. We instructed participants to avoid mixing with urine or water and wash hands with soap after collection. Instructions were given in person in a short letter.

To determinate the presence of trophozoites and other parasite stages, we applied a rapid concentration with saline solution (29) and direct observation method by standard microscopy to analyze fresh stool samples (30). The slide analyses were done by microscopy under 400X with saline solution and lugol, and confirmed with blue methylene-stained stool smear under 1000X (31). All samples were aliquoted into cryovials and stored at −80°C.

### Statistical analysis

Independent associations between *Blastocystis* infection and categorial demographic and sanitary conditions were analyzed with Chi-square and Fisher’s exact test. A multivariable logistic regression model was built to identify adjusted risk factors for *Blastocystis* infection. A linear regression analysis was used to assess the association between humans and animals *Blastocystis* infection in each home. All analyses were performed using R 3.6.2 (32).

## Results

We analyzed stool samples from 337 people and enrolled 232 participants during the campaigns. Participants were classified into 78 index cases, 110 relatives (41 *Blastocystis*-infected and 69 *Blastocystis*-uninfected) and 44 controls who were negative to any intestinal parasite. The age range of participants was 01 to 81 years old (mean=37 SD=21.8), percentage of female participants was 55.6% (n=129). 122 participants are from urban locations, 101 in periurban location and 9 came from rural areas. The education level of participants was 54 elementary school, 44 high school and 128 with college or advanced degrees. Professionally, our sample included 39 homemakers, 74 students, 42 employees, 66 independents contractor and 5 retirees.

We identified seven species of intestinal parasites, including two pathogenic parasites. The prevalence of *Blastocystis* in the study sample was 51.3% with a coinfection percentage of 19%. The second, more prevalent parasite was *Entamoeba coli* (19%) as described in figure 5. To explore the relationship between gastrointestinal symptomatology and *Blastocystis* infection, we included only *Blastocystis* single infection and uninfected participants. Symptomatic participants showed a no significant difference in *Blastocystis* prevalence (41.4%) compared with asymptomatic participants (49.1%). The two most frequent symptoms were flatulence and abdominal pain (44% and 36%), and the less frequent was IBS (5.3%). Statistical analysis (n=167) shows that symptomatology was not significantly different between infected and uninfected *Blastocystis* groups. Specific symptoms also were not significantly associated with *Blastocystis* infection (table 1). We observed a higher percentage of *Blastocystis* infection in male participants across all age groups except under 5 years old and there was a positive trend of infection associated with age (Fig 3).

**Table 1.** Gastrointestinal symptoms of *Blastocystis*-infected and uninfected participants.

| Variable | *Blastocystis* infection |  | *P*-value |
| --- | --- | --- | --- |
|  | Uninfected<br>(n=92) | Infected<br>(n=75) |  |
| Abdominal pain | 36 (39.1%) | 27 (36.0%) | 0.799* |
| Nausea and vomiting | 14 (15.2%) | 11 (14.7%) | 1* |
| Flatulence | 46 (50.0%) | 33 (44.0%) | 0.537* |
| Constipation | 28 (30.4%) | 16 (21.3%) | 0.25* |
| Diarrhea | 27 (29.3%) | 17 (22.7%) | 0.425* |
| IBS | 3 (3.3%) | 4 (5.3%) | 0.702† |

† Fisher \* Chi square

**Figure 3.**
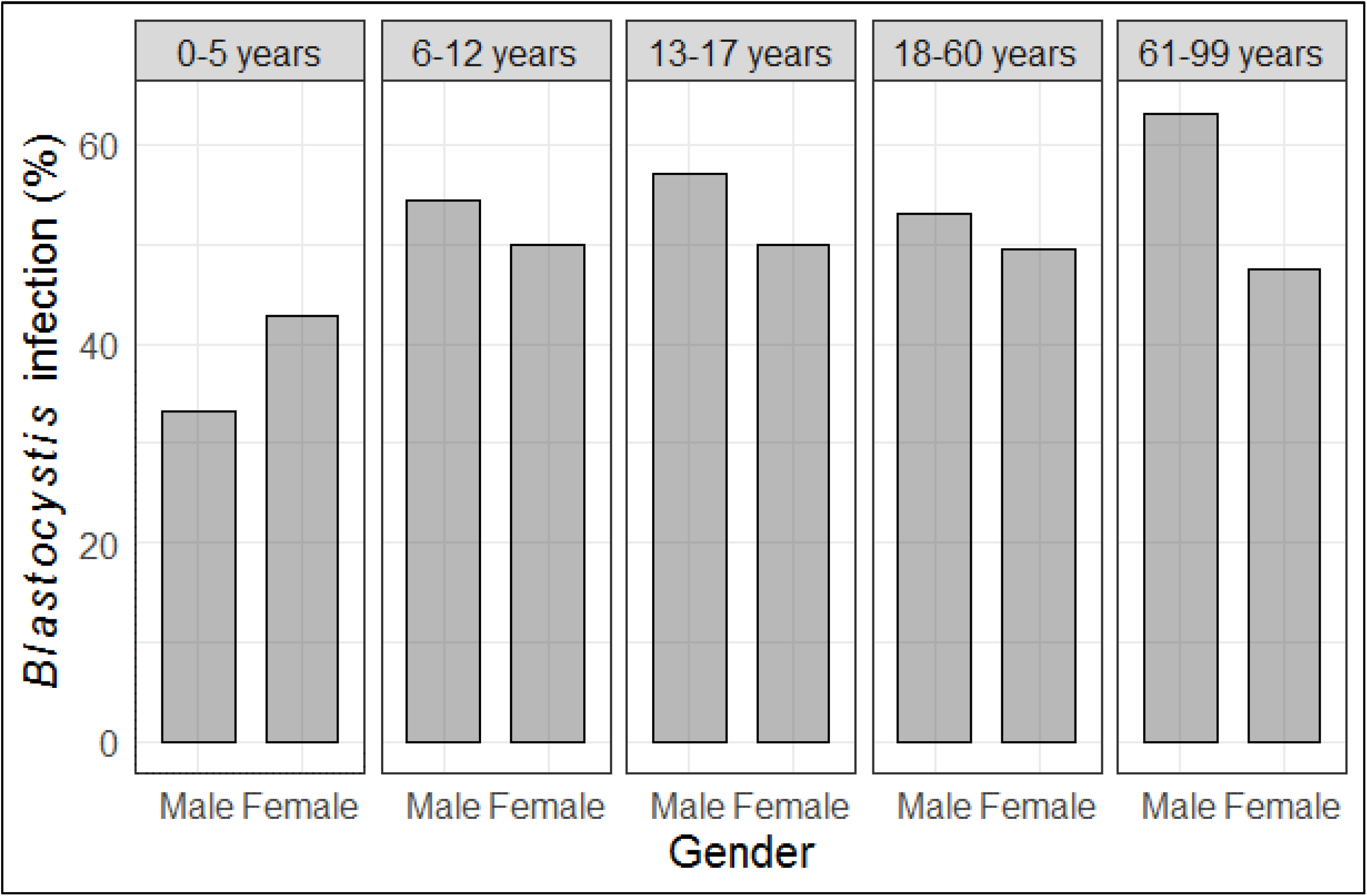
Percentage of *Blastocystis* infection associated to age and location.

**Figure 4.**
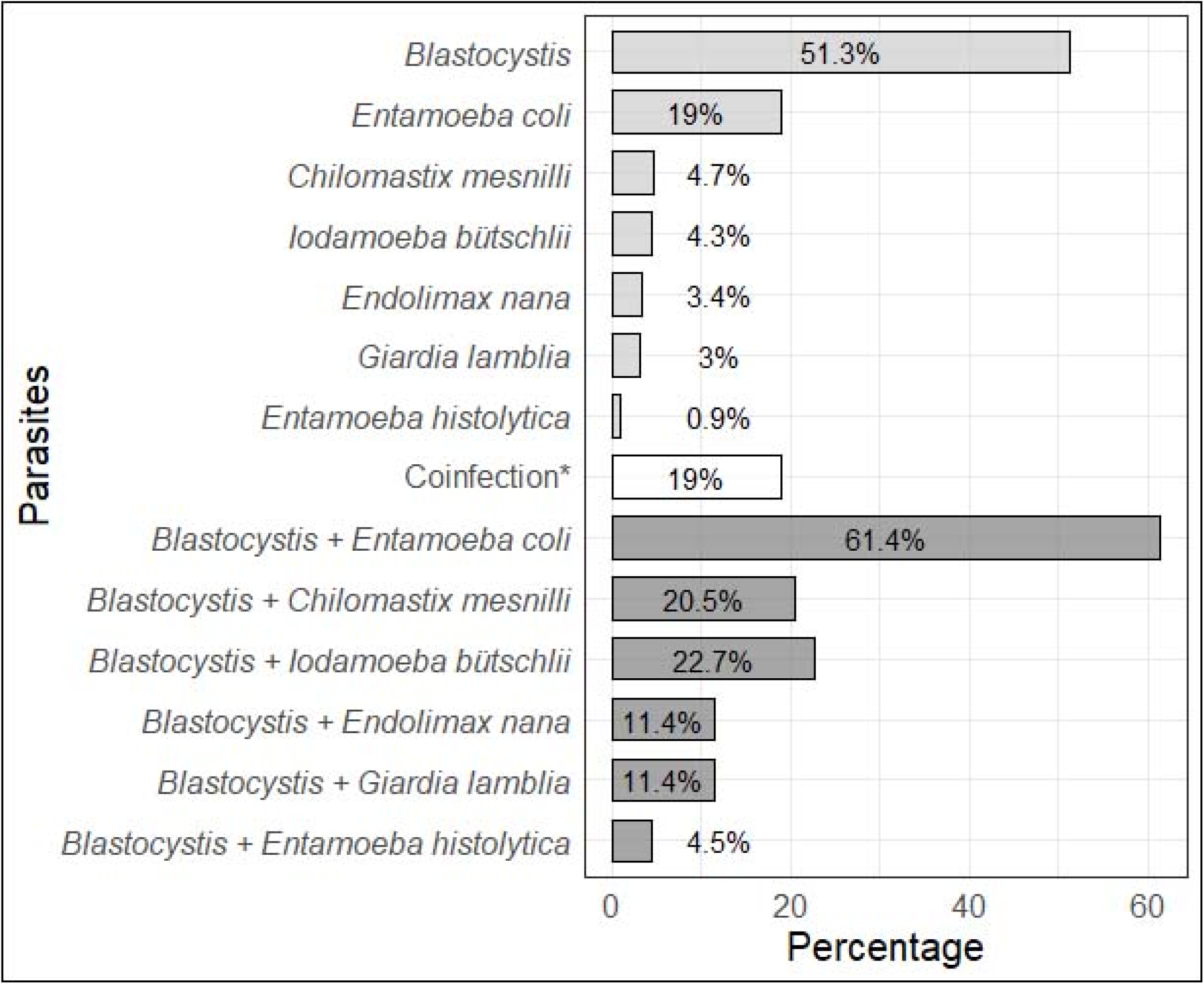
Percentage of intestinal parasites and *Blastocystis* coinfection in the study sample from Arequipa, Peru in 2019 (n=232). *Includes only *Blastocystis* coinfection with other parasites (*Blastocystis* coinfection in dark grey).

To set the statistical analysis for comparisons, we grouped our variables into sanitary variables and demographic variables, that include hygienic habits. The first group includes variables related to sanitary conditions such as location, water supply, body-waste disposal, presence of animals and vectors. For all the analyses we dropped participants from the rural location, and collapsed the water supply category into domiciliary tap water and other water supply that included public standpipe, water tank and water well. Body-waste disposal was collapsed into piped sewer system and latrine, which also included silo (a basic and precarious latrine). Table 2 presents the statistical analysis for independent association of sanitary conditions, which was performed at house level (n=117), including houses from index cases and negative controls. We observed a higher percentage of *Blastocystis* infection in periurban areas as compared to urban areas (p<0.001). Also, there were significant association between *Blastocystis* infection and body-waste disposal, presence of rabbits and rodents. Other variables show no statistical association with *Blastocystis* infection in homes from index cases and negative controls.

**Table 2.** Sanitary characteristics of houses from *Blastocystis*-infected and negative controls

| Variable | *Blastocystis* infection |  | *P*-value |
| --- | --- | --- | --- |
|  | Uninfected<br>n=44 | Infected<br>n=73 |  |
| Location |  |  |  |
| *Urban* | 37 (84.1%) | 31 (42.5%) | **0.0001\*** |
| *Periurban* | 7 (15.9%) | 42 (57.5%) |  |
| Water supply |  |  |  |
| *Domiciliary tap water* | 43 (97.7%) | 66 (90.4%) | 0.255† |
| *Other water supply* | 1 (2.3%) | 7 (9.6%) |  |
| Body-waste disposal |  |  |  |
| *Piped sewer system* | 43 (97.7%) | 63 (86.3%) | **0.051\*** |
| *Latrine* | 1 (2.3%) | 10 (13.7%) |  |
| Presence of dog | 24 (54.5%) | 42 (57.5%) | 0.902* |
| Presence of cat | 10 (22.7%) | 24 (32.9%) | 0.337* |
| Presence of guinea pig | 8 (18.2%) | 17 (23.3%) | 0.675* |
| Presence of rabbit | 1 (2.3%) | 14 (19.2%) | **0.018\*** |
| Presence of poultry | 11 (25.0%) | 18 (24.7%) | 1* |
| Presence of flies | 31 (70.5%) | 50 (68.5%) | 0.987* |
| Presence of cockroach | 6 (13.6%) | 16 (21.9%) | 0.386* |

† Chi square \* Fisher

The second group included variables related to demographic and hygienic habits of participants such as gender, age, education level, economic activity, food consumption place, kind of water consumption, handwashing habits and cleanliness of hands and nails at the time of the survey. The economic activity represents participants’ occupations, where different professions were identified and summarized as students, homemakers, employees, independent contractors and retired. We performed the statistical analysis at individual level (n=155) with *Blastocystis* infected and uninfected participants from *Blastocystis*-positives homes to determinate intra-household factors associated with *Blastocystis* transmission. We do not include the few participants whose responses to hygienic habits questions was “sometimes.” The outcome is presented in table 3, where none variable shows a significant association with *Blastocystis* infection.

**Table 3.** Characteristics of participants at individual-level from *Blastocystis*-infected homes.

| Variable | *Blastocystis* infection |  | *P*-value |
| --- | --- | --- | --- |
|  | Uninfected<br>n=68 | Infected<br>n=87 |  |
| Gender |  |  |  |
| *Female* | 42 (61.8%) | 44 (50.6%) | 0.219* |
| Age |  |  |  |
| 0-5 years | 4 (5.9%) | 4 (4.6%) | 0.815† |
| 6-12 years | 13 (19.1%) | 17 (19.5%) |  |
| 13-17 years | 4 (5.9%) | 6 (6.9%) |  |
| 18-60 years | 41 (60.3%) | 47 (54.0%) |  |
| 61-99 years | 6 (8.8%) | 13 (14.9%) |  |
| Education level |  |  |  |
| *Elementary* | 17 (25.8%) | 21 (24.7%) | 0.918* |
| *High school* | 17 (25.8%) | 20 (23.5%) |  |
| *University* | 32 (48.5%) | 44 (51.8%) |  |
| Economic activity |  |  |  |
| *Student* | 27 (40.9%) | 31 (38.3%) | 0.925* |
| *Homemaker* | 11 (16.7%) | 13 (16.0%) |  |
| *Employees* | 8 (12.1%) | 13 (16.0%) |  |
| *Independent contractor* | 20 (30.3%) | 24 (29.6%) |  |
| Food consumption place |  |  |  |
| *House* | 45 (66.2%) | 61 (70.1%) | 0.636† |
| *Restaurant* | 1 (1.5%) | 0 - |  |
| *House and restaurant* | 15 (22.1%) | 20 (23.0%) |  |
| *House and kiosk* | 7 (10.3%) | 6 (6.9%) |  |
| Kind of water consumption |  |  |  |
| *Boiled water* | 54 (79.4%) | 68 (78.2%) | 1† |
| *Tap water* | 4 (5.9%) | 6 (6.9%) |  |
| *Both* | 10 (14.7%) | 13 (14.9%) |  |
| Don't wash hands after using bathroom | 6 (8.8%) | 7 (8.0%) | 1* |
| Don't wash hands before cooking | 13 (19.1%) | 22 (25.3%) | 0.4727* |
| Don't wash hands before eating | 11 (16.7%) | 9 (10.5%) | 0.3794* |
| Don't wash hands after touching animals | 21 (31.3%) | 31 (35.6%) | 0.6994* |
| Had dirty hands at time of survey | 13 (19.1%) | 21 (24.1%) | 0.5796* |
| Had dirty nails at time of survey | 21 (30.9%) | 32 (36.8%) | 0.55* |

† Fisher \* Chi square

**Table 4.** Bivariate and multivariate analysis of *Blastocystis* infection risk factors

| Variable | Bivariate logistic regression |  |  | Multivariate logistic regression |  |  |
| --- | --- | --- | --- | --- | --- | --- |
|  | OR<sup>a</sup> | 95% CI | p value | OR<sup>b</sup> | 95% CI | p value |
| Periurban location | 7.2 | 3.0 - 19.5 | **<0.001** | 3.7 | 1.4 - 10.8 | **0.012** |
| Water supply: Other water supply | 4.6 | 0.8 - 86.8 | 0.101 | 19.3 | 1.5 - 425.2 | **0.038** |
| Body-waste disposal: latrine | 6.8 | 1.2 - 127.6 | **0.024** | 0.1 | 0 - 0.8 | **0.040** |
| Presence of dog | 1.1 | 0.5 - 2.4 | 0.752 | 10.2 | 2.0 - 59.7 | **0.006** |
| Presence of cat | 1.7 | 0.7 - 4.1 | 0.236 |  |  |  |
| Presence of guinea pig | 1.4 | 0.6 - 3.7 | 0.511 |  |  |  |
| Presence of rabbit | 10.2 | 1.9 - 188.4 | **0.003** | 2.0 | 0.6 - 7.3 | 0.258 |
| Presence of poultry | 1.0 | 0.4 - 2.4 | 0.967 |  |  |  |
| Presence of flies | 0.9 | 0.4 - 2.0 | 0.824 |  |  |  |
| Presence of cockroach | 1.8 | 0.7 - 5.3 | 0.258 |  |  |  |
| Female participants | 0.6 | 0.3 - 1.2 | 0.163 | 0.6 | 0.3 - 1.3 | 0.178 |
| Age group |  |  |  |  |  |  |
| 13-17 years | 1.5 | 0.2 - 10.4 | 0.672 |  |  |  |
| 18-60 years | 1.1 | 0.3 - 5.1 | 0.853 |  |  |  |
| 61-99 years | 2.2 | 0.4 - 12.4 | 0.370 |  |  |  |
| Education |  |  |  |  |  |  |
| High school | 1.0 | 0.4 - 2.4 | 0.916 |  |  |  |
| University | 1.1 | 0.5 - 2.4 | 0.789 |  |  |  |
| Food consumption place |  |  |  |  |  |  |
| House and restaurant | 1.0 | 0.5 - 2.2 | 0.967 |  |  |  |
| House and kiosk | 0.6 | 0.2 - 2.0 | 0.437 |  |  |  |
| Kind of water consumption |  |  |  |  |  |  |
| Tap water | 1.2 | 0.3 - 4.9 | 0.794 |  |  |  |
| Boiled and tap water | 1.0 | 0.4 - 2.6 | 0.945 |  |  |  |
| Don't wash hands after using bathroom | 1.1 | 0.3 - 3.5 | 0.863 |  |  |  |
| Don't wash hands before cooking | 0.7 | 0.3 - 1.5 | 0.359 |  |  |  |
| Don't wash hands before eating | 1.7 | 0.7 - 4.5 | 0.265 |  |  |  |
| Don't wash hands after touching animals | 0.8 | 0.4 - 1.6 | 0.576 |  |  |  |
| Had dirty hands at time of survey | 1.3 | 0.6 - 3 | 0.452 |  |  |  |
| Had dirty nails at time of survey | 1.3 | 0.7 - 2.6 | 0.441 |  |  |  |

<sup>a</sup>Unadjusted OR values; <sup>b</sup>Adjusted OR values

The second group included variables related to demographic and hygienic habits of participants such as gender, age, education level, economic activity, food consumption place, kind of water consumption, handwashing habits and cleanliness of hands and nails at the time of the survey. The economic activity represents participants’ occupations, where different professions were identified and summarized as students, homemakers, employees, independent contractors and retired. We performed the statistical analysis at individual level (n=155) with *Blastocystis* infected and uninfected participants from *Blastocystis*-positives homes to determinate intra-household factors associated with *Blastocystis* transmission. We do not include the few participants whose responses to hygienic habits questions was “sometimes.” The outcome is presented in table 3, where none variable shows a significant association with *Blastocystis* infection.

Bivariate analyses highlighted that location (OR=7.2), other water supply (OR=4.6), use of latrine (OR=6.8) and presence of rabbits (OR=10.2) were statistically associated with *Blastocystis* infection. To set up the multivariate analysis, we included variables yielding a p-value less than or equal to 0.2 in the bivariate analysis and performed a stepwise approach to determinate interaction and effect of independent variables on *Blastocystis* infection. Multivariate analysis showed that the only variables that affect *Blastocystis* infection were living in periurban areas (OR=3.7), having or using a different water supply than domiciliary tap water (OR=19.3) and presence of dogs (OR=10.2); the most significant association was with domiciliary water supply. Other variables in the analysis showed no significant association with *Blastocystis* infection, mainly variables related to demographic and hygienic habits.

## Discussion

The controversy surrounding *Blastocystis* in recent decades has resulted in increased interest in developing detailed studies focused on determining whether or not the parasite is pathogenic or beneficial to infected humans (9,33). Most previous studies in Arequipa focused on reporting intestinal parasites’ prevalence in elementary school children. Those reports identified *Blastocystis* as a recurrent parasite in this population, though they did not link epidemiological data to clinical outcomes. This is the first study that aimed to use the One Health approach to identify social determinants and individual factors associated with *Blastocystis* infection in Arequipa, Peru.

The prevalence of *Blastocystis* infection found in our study was similar to other values reported in previous studies in Peru (22–24). However, it was in the lower boundary of the range previously reported in Arequipa (48.29%-81.9%) (25,26). This lower prevalence can be explained by the fact that previous studies in Arequipa were focused on school children and we included all age groups. We observed prevalence of *Blastocystis* infection above 50% in school children and lower values in people ages 60 years or older and infants. However, *Blastocystis* was the most prevalent parasite in this study sample, which is similar to other studies around the world (34,35). Likewise, we found no significant difference in the infection rate according to age or sex, a trend which has been reported in other studies (36,37). This finding counters findings from rural populations of Brazil, where *Blastocystis* infection was linked to being female and being a homemakers (15).

This study highlights living in periurban communities as one of the most important risk factors for the transmission of *Blastocystis* in humans. This is supported by a WHO report on social determinants of health (38), which described how periurban areas often have unhealthy sanitary conditions linked to poverty that facilitate health problems and increase rates of infectious diseases. These conditions are quite similar to rural areas, the origin of local migration to periurban areas of Arequipa (39).

We identified that water supply is the other main risk factor for *Blastocystis* transmission in periurban locations. Due to the limited access to domiciliary tap water, people in periurban areas often consume water from standpipes, water tanks or wells. Some studies have reported similar results in areas with poor hygiene and sanitation and facilities with a high prevalence of *Blastocystis* (14,16) whereas others studies did not find this association (19). *Blastocystis* has been reported in bodies of water in the veterinary literature (16,40) and in contaminated water, which has been reported as a source of infection (13).

The presence of animals is a common characteristic in periurban location in Arequipa, where community members raise animals as a food resource, a rural custom brought to city due to migration (39). We find that the presence of dogs and rabbits is associated with *Blastocystis* infection in humans. This finding suggests that animals can play a key role in *Blastocystis* zoonotic transmission into infected homes and act as secondary reservoirs for *Blastocystis* (41). Therefore, a One Health approach may be helpful in understanding the epidemiology of *Blastocystis*. This approach is also supported by previous studies that reported the occurrence of *Blastocystis* infection in different animal species like poultry, cattle, pigs and dogs (42,43). Similar associations to these animal species have been reported in endemic zoonotic disease in Arequipa (44).

This study provides an overview socio-demographic determinant associated with *Blastocystis* infections; however, we could not determinate the commensal or pathogenic role of *Blastocystis* in humans. Some limitations of our study are the different procedures for participant recruitment and low participation in our campaigns. We were also unable to enroll all household members of every index case to estimate the intra-household transmission rate. Furthermore, our study design was cross-sectional; therefore, it is possible that *Blastocystis* infection is transient and with a high rate of coinfection. Finally, the sampling we used for the university-based campaign was not random and some of the variables we analyzed are correlated. Future studies would benefit from improved recruitment strategies, including rural areas, and conducting longitudinal studies in humans and their animals to assess changes in *Blastocystis* infection status and potential transmission pathways.

## Conclusions

We found that human *Blastocystis* infection is associated with a group of factors that are found in periurban environments in the city of Arequipa, such as using an alternative not domiciliary water supply and using latrines for human waste body-waste disposal. The role of animals like dogs and rabbits further elucidated to understand if they act as reservoirs or recipients of the parasite (zooanthroponosis). We did not find a link between *Blastocystis* infection and gastrointestinal symptomatology in our study.

## Data Availability

The data will be available if requested by email to the first author

## Acknowledgments

Thanks to Dr. Irmia Paz (UNSA) for her revision and suggestions and Julianna Shinnick for her valuable support and advise related to English review and Mr. Cirilo Neyra for technical support. We would like to give a special acknowledgement to principals from the schools that participate in the study, for their enthusiasm and support to participate in the study looking for the welfare of their students:

To Berardo Jesus Carrasco Castro. Principal at Sagrado Corazón de Jesus 40078 Primary school from Tio Chico, Sachaca.

To Salome Eliana Alvarez Marquina. Principal at Tío Chico Kinder garden school from Tio Chico, Sachaca.

To Deysi Lilian Esquiagola Tapia, Principal at Karol Jozef Wojtyla, Primary school from Socabaya.

To Principal at Jose Carlos Echavarry Osacar, Primary school from Tiabaya.

